# Sex-Based Differences in Long-Term Lipid Metabolism, Inflammation and Stress Regulation After Non-Severe Paediatric Burns

**DOI:** 10.1101/2025.09.23.25336456

**Authors:** Monique J Ryan, Eva Kierath, Samantha Lodge, Reika Masuda, Jacqueline Davis, Nina D’Vaz, Lucy W Barrett, Nicola Gray, Fiona M Wood, Elaine Holmes, Mark W Fear, Luke Whiley

## Abstract

Paediatric burn injuries are a global health concern with long-term health consequences, such as psychological, immune, and cardiovascular complications, that can persist even after non-severe injuries. Emerging evidence suggests that biological sex may influence post-burn outcomes in children, as female burn survivors have been shown to experience higher mortality, scarring, anxiety, depression, and poorer quality of life compared to males. This study addresses a critical research gap by examining sex-specific lipidomic and inflammatory responses following paediatric non-severe burn injury. Children under five years were recruited as a part of the Childhood Burn Injury Biobank at hospital admission with longitudinal follow-ups and non-burn controls aged 1 and 3 were collected from the ORIGINS cohort. Plasma lipid profiling and acute-phase glycoprotein systemic inflammatory markers (GlycA and GlycB) were quantified using metabolic phenotyping with hair cortisol provided as a longitudinal measure of stress. Lipidomic analysis revealed acute-phase disruptions in fatty acids and lysophospholipids in both sexes but only females demonstrated a persistent increase of arachidonic acid (FA 20:4) and depletion of monoacylglycerols more than a year post-injury. Females also had significantly higher acute-phase GlycA/GlycB levels around the time of injury and exhibited increasing variability in hair cortisol over time, while male burn survivors did not. These findings highlight a sex-specific response between lipid metabolism, systemic inflammation and stress in paediatric recovery from non-severe burns. Understanding these differences may guide the development of targeted psychological and physiological strategies to improve long-term outcomes for young burn survivors.

## Introduction

Paediatric burn injuries are a significant global health burden. In Western Australia alone, children under the age of 15 account for approximately 35% of all burn-related hospital admissions, which is a trend that is reflected across both high- and lower-middle-income countries^1^. The consequences of burn trauma in children extend well beyond the initial injury. Paediatric burn traumas can lead to long-term cardiovascular, immune, musculoskeletal and nervous system complications, which can persist for decades post-injury even in non-severe cases^2–7^. These long-term complications are often combined with chronic pain, reduced exercise capacity, muscle weakness, scarring, and psychosocial challenges, significantly impacting the quality of life of childhood burn survivors and their families^7^.

Despite improvements in acute burn care, recent research has identified biological sex as an additional risk factor for some of the long-term complications associated with burn injury. In adult cohorts, females have been reported to have twice the mortality rate compared to males when adjusted for total body surface area percentage (TBSA%) and age^8–12^. In paediatric cases, this is reflected with female children experiencing higher rates of mortality following severe burn injuries^13^, higher rates of pathologic scarring^14,15^ and poorer long-term psychological outcomes, including greater incidence of anxiety, depression, with female burn survivors reporting a lower overall quality of life^16,17^. However, there are discrepancies in the literature, with some studies finding that being female was protective of adverse burn outcomes, and others reporting no significant sex differences in terms of patient outcomes^18–21^. These inconsistencies, coupled with limited research, highlight the need for further studies to better understand the impact of sex on paediatric burn injury recovery and long-term outcomes.

A potential mechanistic area for sex-differences in outcomes after paediatric burn injury may lie between stress, inflammation and metabolism, particularly lipid-mediated inflammation. Inflammation and lipid metabolism are two correlated biological systems that are known to influence each other^22^ and contribute to the pathophysiology of various diseases, including cancer^23^, cardiovascular disease^24^, diabetes^25^ and even burn trauma^26^. Studies in other chronic inflammatory conditions, such as asthma, cystic fibrosis and sickle cell anaemia, have shown that females under ten exhibit a stronger increase in inflammatory markers compared to males^27,28^. These studies identified higher neutrophil counts and C-reactive protein levels with more frequent or severe clinical symptoms in females^27,28^, which may also be relevant in the context of recovery after paediatric burn injuries.

Psychological stress is another factor known to modulate inflammation through steroid hormones such as cortisol^29,30^ and has recently been associated with lipid metabolic changes in children^29^. Research into childhood pain responses has found that young females are more vulnerable to long-term anxiety and stress compared to males^31^, with adult females who experienced childhood burn trauma reporting significantly higher rates of psychological adversity^16^. Furthermore, studies on childhood trauma and stress have identified a higher risk of developing metabolic diseases, such as obesity and cardiovascular conditions, in women later in life. These outcomes are partly attributed to elevated cortisol levels and dysregulation of the hypothalamic-pituitary-adrenal (HPA) axis^32,33^. Together, this evidence points to a potentially underexplored, systemically linked triad of stress, inflammation, and metabolism that may influence sex-based differences in responses to paediatric burn injury, both in the short and long term.

Recent research has identified sex differences in the lipidome in both healthy adults and children ^34–37^. In addition to this sex-difference in the lipidome, lipid-related inflammatory pathways may provide insights into sex-specific responses during burn injury and recovery. While our previous pilot study identified lipidomic alterations up to three years post-injury in paediatric burn survivors^38^, sex differences in these lipidomic responses have not yet been explored. The current study seeks to address this gap by conducting a detailed metabolic phenotyping analysis in children under the age of five, to characterise lipid-related inflammatory and cortisol differences between male and female children following non-severe burn trauma. Understanding these sex-specific molecular pathways may help inform more targeted and effective interventions to improve long-term outcomes for all paediatric burn survivors.

## Methods

### Study design

The Childhood Burn Injury Biobank recruited children (n=237) from Perth Children’s Hospital who were treated for a burn injury of any severity and were less than 16 years old, with informed consent granted through a parent or guardian. For this study, only those patients that were ≤5 years of age at time of injury were included (n=118). Ethics for this biobank and subsequent research was approved by The Child and Adolescent Health Service Ethics Committee (Ethics No. RGS0000001403) and by Murdoch University Human Research Ethics Committee (Ethics No. 2021/160) in Western Australia.

For non-burn controls, samples from children aged 1 and 3 (n=299) were provided by ORIGINS Biobank^39,40^. ORIGINS is a population cohort in Western Australia that follows 10 000 children from birth until the age of 10. Ethics for use of these samples was approved by the Ramsay Health Care WA / SA Human Research Ethics Committee (Reference: 2146W).

### Plasma collection

Plasma was collected in lithium heparin collection tubes using matched protocols by both the Childhood Injury Biobank and ORIGINS for each child^40^. Non-burn controls had one sample taken at ages 1 and 3. For the paediatric burn cohort, blood was drawn on admission to the Burns Unit when receiving treatment for a burn injury. Follow-up samples were collected on re-visit to hospital, which fell into the following time categories: ‘Acute phase’ which was <14 days from the initial injury, ‘Recovering phase’ between 14 days to 1-year post-admission and once deemed ‘Recovered’, over a year post-admission. Ethics approval for the analysis of these samples was obtained from Murdoch University Human Research Ethics Committee (Ethics No. 2021/160).

### Hair cortisol measurements

Hair strands with ∼5mm diameter were collected from the posterior vertex region of the scalp. The hair tip was placed into a 2mL cryotube and cut near the scalp to ensure cortisol measurements closest to time since injury. Only 3 cm from the scalp (roughly 3 months of growth) was used for analysis. Hair strands were collected up to 1 week post-admission to hospital (to mimic a pre-burn injury baseline), between 1 week to 1 month, 3 to 6 months, 6 to 12 months and over 1 year since injury.

Cortisol extraction from the hair and subsequent analysis was performed by Stratech Scientific (Mona Vale, Australia) according to their validated protocol. In brief, hair strands were weighed prior to extraction for correction post-analysis, then mechanically crushed and extracted using 100% methanol. Samples were dried down to remove the solvent before reconstitution in phosphate-buffered saline. Cortisol was analysed using a salivary cortisol enzyme-linked immunosorbent assay kit (Salimetrics, Pennsylvania, USA) with a calibration range of 0.012 - 3 µg/dL.

### Comprehensive lipid profiling

Lipid profiling on the plasma samples was performed in accordance with an established protocol that has previously been published and validated^41^. In brief, 20 µL of plasma was extracted with 180 µL of isopropanol (Thermo Fisher Scientific, Malaga, Western Australia, Australia) containing Ultimate SPLASH™ ONE, SphingoSPLASH™ I, 17:1 Lyso PS and 18:1-d7 MG from Avanti Polar Lipids (Sigma-Aldrich, North Ryde, New South Wales, Australia), and Arachidonic acid-d5, Linoleic acid-d11, Oleic acid-d9, Palmitic acid-d5 and Stearic acid-d4 from Cayman Chemical (Sapphire Bioscience, Redfern, New South Wales, Australia). Samples were vortexed, chilled at −20 °C and centrifuged before supernatant extraction (50 µL) into 96-well plates for liquid chromatography tandem mass spectrometry (LC-MS/MS) analysis. For detection of lipids from hair strands, 20 µL of the leftover methanol extraction for the cortisol measurements underwent the same procedure as in the described plasma protocol. A list of internal standards and their concentrations are available in Supplementary Table S1. LC-MS/MS was performed using a SCIEX ExionLC and QTRAP 6500+ mass spectrometer to detect 1163 lipid species and pooled quality control (QC) samples created from combining all cohort samples were run intermittently to assess instrument performance.

Raw spectra were imported into Skyline v24.1 (MacCoss, WA, USA) for peak integration, while data pre-processing and filtering were carried out in R (v4.4.3, R Foundation, Vienna, Austria) using RStudio IDE (build 2024.12.1+563, RStudio, Boston, MA, USA)^42^. Lipid species and samples with >50% missing values were excluded. For those with <50% missing values, missing data were imputed using half of the minimum detected value for each lipid. Lipid species with a relative standard deviation >30% in pooled QC samples were also removed. To correct for signal drift, each lipid concentration was adjusted using the Random Forest method within the *statTarget* R package (available from github.com/statTarget) using pooled QCs as reference.

### 1H NMR spectroscopy

Plasma was prepared for ^1^H NMR spectroscopy using an established validated protocol^43–45^. In summary, a 600 MHz Bruker Avance III HD spectrometer coupled with a BBI probe and SampleJet robot (Bruker BioSpin, Massachusetts, USA) was employed to analyse plasma samples maintained at 5 °C in the system. Plasma was diluted 1:1 with a phosphate buffer (75 mM Na_2_HPO_4_, 2 mM NaN_3_, and 4.6 mM sodium trimethylsilyl propionate-[2,2,3,3-2H_4_] (TSP) in D_2_O, pH 7.4 ± 0.1) before 180 µL of the mixture was added into 3 mm SampleJet NMR tubes. ^1^H NMR spectroscopy was performed per Bruker *in vitro* Diagnostics research (IVDr) methods for ^1^H 1D and PGPE experiments^44,46^. ^1^H 1D experiments with solvent pre-saturation ran for an experiment time of 14 min 53 s acquiring 128 scans, 98 K data points with a spectral width of 30 ppm, followed by the PGPE experiment run for 16 min and 47 s obtaining 256 scans, 98 K data points and a spectral width of 30 ppm. The PGPE experiment was run with a secondary irradiation field for solvent suppression (set at the water frequency) to ensure the capture of *N*-glycan inflammatory markers GlycA (arising from *N*-acetylneuraminic acid moieties) and GlycB (originating from *N*-acetylglucosamine moieties) within the spectra.

Data pre-processing was performed in R (v4.4.3) using in-house and open-source packages *nmr-parser* (available from github.com/phenological/nmr-parser) and *nmr-spectra-processing* (available from github.com/phenological/nmr-spectra-processing). Spectra obtained from the PGPE experiment was corrected for instrument variability using the ERECTIC factor and baseline corrected with an asymmetric least-squares routine. Signals from acetylated glycoproteins known to be associated with inflammation^47,48^, GlycA and GlycB, were integrated by summing fixed spectral regions at δ 2.05−2.09 (GlycA) and δ 2.09−2.12 (GlycB).

### Statistical analysis and data visualisation

Statistical analysis and visualisation of the data was conducted using R (v4.4.3) and the RStudio IDE (build 2024.12.1+563). Mann-Whitney U tests were applied for univariate statistical analysis with significance set at *p*-value < 0.05. Multivariate analysis including orthogonal projections to latent structures-discriminate analysis (OPLS-DA) was performed using the *mva.plots* R package (v0.0.8) (available from github.com/phenological/mva-plots)and eruption plots were generated for each of the models. Eruption plots, the multivariate equivalent of a volcano plot^49^, enable the visualisation of the univariate effect size through Cliff’s delta (x-axis) combined with the multivariate OPLS-DA predictive loadings (y-axis). Lipid subclasses were calculated by summing all the lipid species within each subclass. There is an exception with triacylglycerols (TGs) where after summing the subclass is divided by three due to the limitation in the LC-MS/MS analysing the same TG species three times for each sidechain^50^. All visualisations utilising data from other packages were produced through *ggplot2* from github.com/tidyverse/ggplot2.

## Results

### Clinical outcomes of sex in paediatric patients

For the overall paediatric cohort, there were no statistically significant differences between females and males in terms of age, Fitzpatrick skin type, burn depth, burn source, wound and scar management, or surgical requirements (Table 1). Females had a significantly longer hospital stay (*p*-value = 0.043) and a higher incidence of face, neck and forearm burns (*p*-value <0.01) compared to males. For the overall cohort, both males and females had <10% TBSA which is classified as a non-severe burn.

**Table 1.** Demographics of the total paediatric burn cohort split by sex and corresponding *p*-values. Significant *p*-values (<0.05) are bolded.

| Characteristic | Overall, N = 118<sup>1</sup> | Female, N = 51<sup>1</sup> | Male, N = 67<sup>1</sup> | *p*-value<sup>2</sup> |
| --- | --- | --- | --- | --- |
| Age at injury | 1.76 (1.30, 2.92) | 1.78 (1.39, 2.99) | 1.74 (1.25, 2.87) | 0.5 |
| **Fitzpatrick score** |  |  |  | 0.6 |
| *I* | 14 (12%) | 4 (7.8%) | 10 (15%) |  |
| *II* | 62 (53%) | 26 (51%) | 36 (54%) |  |
| *III* | 17 (14%) | 10 (20%) | 7 (10%) |  |
| *IV* | 15 (13%) | 6 (12%) | 9 (13%) |  |
| *V* | 5 (4.2%) | 2 (3.9%) | 3 (4.5%) |  |
| *VI* | 5 (4.2%) | 3 (5.9%) | 2 (3.0%) |  |
| **Aboriginal status** |  |  |  | 0.4 |
| *Aboriginal* | 15 (13%) | 5 (9.8%) | 10 (15%) |  |
| *Other* | 102 (87%) | 46 (90%) | 56 (85%) |  |
| **TBSA%** | 2.0 (1.0, 5.4) | 3.0 (1.0, 5.5) | 2.0 (1.0, 5.0) | 0.2 |
| **Burn depth** |  |  |  | >0.9 |
| *Deep* | 49 (42%) | 20 (41%) | 29 (43%) |  |
| *Mid dermal / partial thickness* | 45 (39%) | 19 (39%) | 26 (39%) |  |
| *Full thickness* | 20 (17%) | 9 (18%) | 11 (16%) |  |
| *Superficial partial* | 2 (1.7%) | 1 (2.0%) | 1 (1.5%) |  |
| **Burn source** |  |  |  | >0.9 |
| *Scald* | 62 (53%) | 28 (56%) | 34 (51%) |  |
| *Contact* | 36 (31%) | 14 (28%) | 22 (33%) |  |
| *Friction* | 14 (12%) | 6 (12%) | 8 (12%) |  |
| *Flame* | 3 (2.6%) | 1 (2.0%) | 2 (3.0%) |  |

| Characteristic | Overall, N = 118<sup>1</sup> | Female, N = 51<sup>1</sup> | Male, N = 67<sup>1</sup> | p-value<sup>2</sup> |
| --- | --- | --- | --- | --- |
| *Electrical* | 2 (1.7%) | 1 (2.0%) | 1 (1.5%) |  |
| **Burn site<sup>3</sup>** |  |  |  |  |
| *Face and neck* | 16 (14%) | 12 (24%) | 4 (6.0%) | **0.006** |
| *Torso* | 40 (34%) | 21 (41%) | 19 (28%) | 0.15 |
| *Arms* | 35 (30%) | 23 (45%) | 12 (18%) | **0.001** |
| *Hands* | 49 (42%) | 25 (49%) | 24 (36%) | 0.15 |
| *Legs* | 24 (20%) | 14 (27%) | 10 (15%) | 0.094 |
| *Feet* | 16 (14%) | 4 (7.8%) | 12 (18%) | 0.11 |
| *Genitalia* | 1 (0.8%) | 0 (0%) | 1 (1.5%) | >0.9 |
| *Buttocks* | 6 (5.1%) | 3 (5.9%) | 3 (4.5%) | >0.9 |
| **Surgery type** |  |  |  | 0.4 |
| *Recell* | 76 (70%) | 31 (65%) | 45 (74%) |  |
| *Split skin graft and Recell* | 18 (17%) | 8 (17%) | 10 (16%) |  |
| *Split thickness skin graft* | 9 (8.3%) | 4 (8.3%) | 5 (8.2%) |  |
| *Biobrane* | 1 (0.9%) | 1 (2.1%) | 0 (0%) |  |
| *Other* | 5 (4.6%) | 4 (8.3%) | 1 (1.6%) |  |
| **Hospital length of stay** | 3 (1, 6) | 4 (2, 8) | 2 (1, 5) | **0.043** |
| **First aid prior to admission** |  |  |  | 0.4 |
| *Correct first aid<sup>4</sup>* | 80 (68%) | 33 (65%) | 47 (70%) |  |
| *Inadequate first aid* | 12 (10%) | 5 (9.8%) | 7 (10%) |  |
| *No first aid within 3 hours* | 4 (3.4%) | 1 (2.0%) | 3 (4.5%) |  |
| *Other cooling measures* | 2 (1.7%) | 0 (0%) | 2 (3.0%) |  |
| *Not stated* | 20 (17%) | 12 (24%) | 8 (12%) |  |
| **Days to re-epithelialisation** | 18 (14, 28) | 18 (15, 32) | 18 (14, 27) | 0.8 |
| **Wound complications** |  |  |  |  |
| *Delayed healing* | 81 (69%) | 33 (65%) | 48 (72%) | 0.4 |
| *Wound infection* | 4 (3.4%) | 3 (5.9%) | 1 (1.5%) | 0.3 |
| **Scar management required** |  |  |  |  |
| *Pressure garment* | 35 (45%) | 17 (45%) | 18 (45%) | >0.9 |
| *Laser* | 23 (29%) | 13 (34%) | 10 (25%) | 0.4 |
| *Silicone application* | 42 (54%) | 20 (53%) | 22 (55%) | 0.8 |
| *Splint addition* | 7 (9.0%) | 3 (7.9%) | 4 (10%) | >0.9 |

<sup>1</sup> Median (IQR); n (%)
<sup>2</sup> Wilcoxon rank sum test; Fisher's exact test; Pearson's Chi-squared test
<sup>3</sup> Note: The same patient may have burns across multiple body sites.
<sup>4</sup> Refers to holding the injury under cool running water for at least 20 minutes<sup>51</sup>.

### Burn healing trajectories are different for males and females after non-severe burn injury when compared to non-burn controls

Both male and female paediatric burn patients displayed distinct lipidomic signatures during the acute phase (<14 days post-injury) of the non-severe burn injury compared to non-burn controls at 1 year of age, which is a similar age to the burn cohort at the time of injury (Figure 1A-B). The OPLS-DA models revealed strong and significant lipidomic differences for males (Figure 1A: R^2^X(cum) = 0.474, R^2^Y(cum) = 0.846, Q^2^(cum) = 0.676) and females (Figure 1B: R^2^X(cum) = 0.575, R^2^Y(cum) = 0.908, Q^2^(cum) = 0.712). Due to the large number of TG lipid species detected (50% of the total lipid species: n=444, total lipid species n = 888), eruption plots (Figure 2) were generated with the lipid subclasses instead to highlight the lipidomic changes that may be otherwise be overshadowed by TGs. The results showed that the lipidomic signatures in males and females were similar compared to non-burn controls in the acute phase (Figure 2A-B). Burn lipidomic signatures were characterised by increases in the concentration of the plasma fatty acid (FA) subclass and decreases in the lysophosphatidylglycerol (LPG) subclass. For males, the increases in FA were a more significant contributor to the model than for females during the acute phase of burn injury.

**Figure 1.**
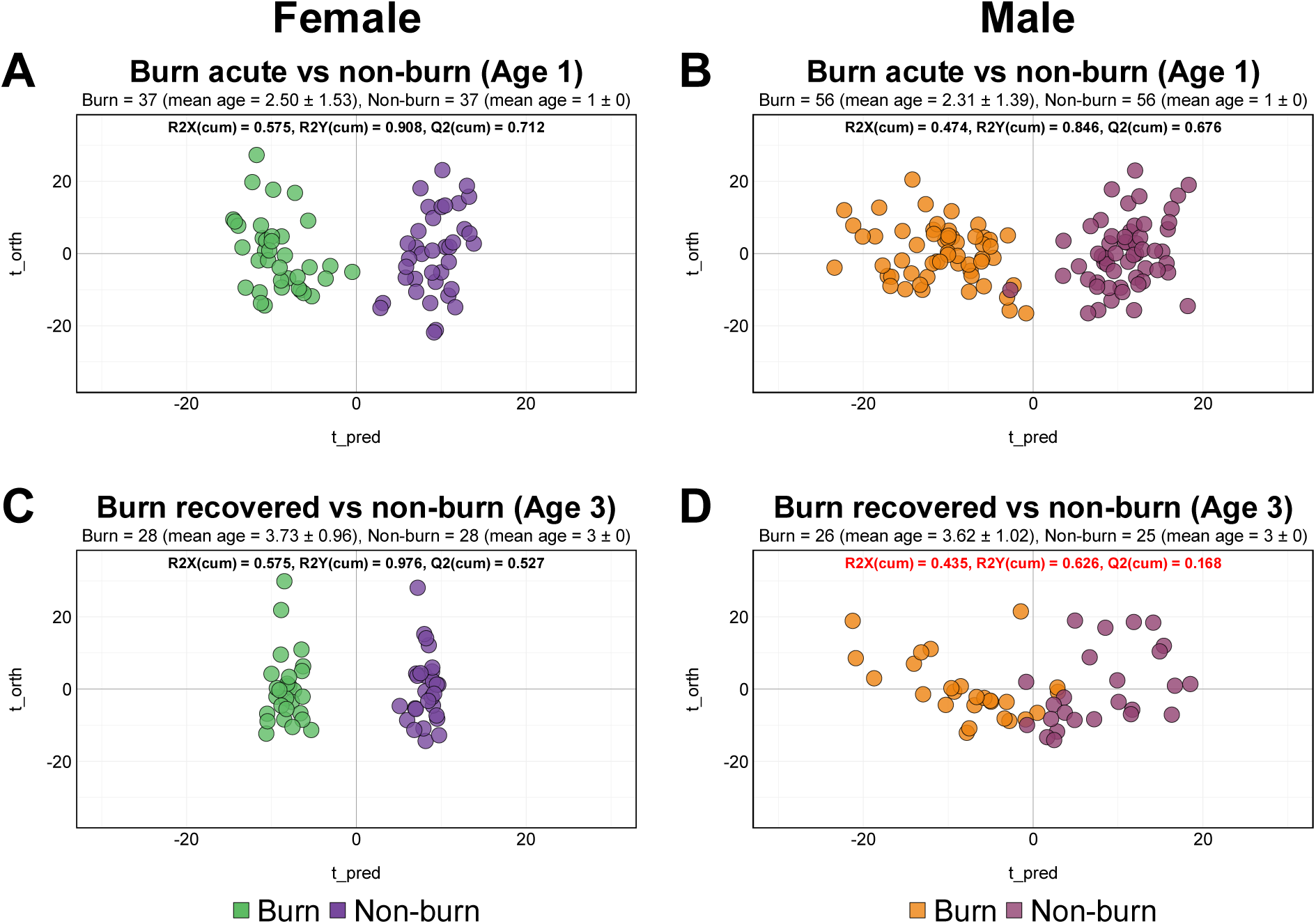
Lipidomic differences between males and females post-non-severe burn injury compared to non-burn controls. OPLS-DA classed by injury are separated in columns by sex (Left: female burn injury = green, female non-burn = purple; Right: male burn injury = orange, male non-burn = burgundy) **A)** Non-burn controls at the age of one compared to the burn acute phase (<14 day from injury) for females, **B)** Non-burn controls at the age of one compared to the burn acute phase (<14 day from injury) for males, **C)** Non-burn controls at the age of three compared to burn cohort when considered ‘recovered’ > 1 year from injury) for females, and **D)** Non-burn controls at the age of three compared to burn cohort when considered ‘recovered’ (> 1 year from injury) for males. *Note:* Metadata provided for the non-burn controls stipulated ages 1 or 3, resulting in a standard deviation of 0 in the plot subtitles.

**Figure 2.**
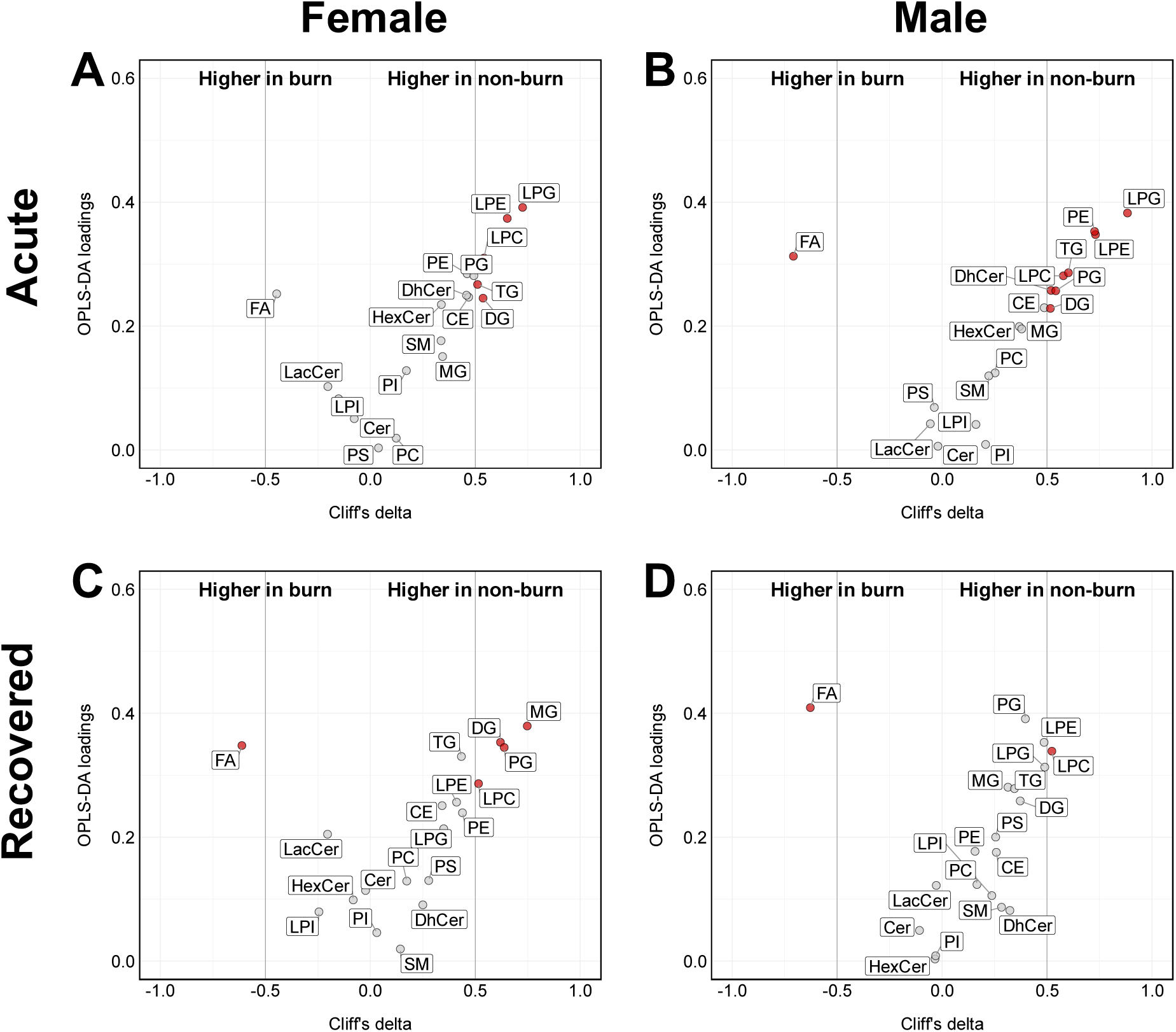
Changes in lipid subclasses over time in females and males within the paediatric burn cohort. Visualised by an eruption plot depicting Cliff’s delta (x-axis) and OPLS-DA predictive loadings (y-axis) of lipid classes subclass coloured by significance (Mann-Whitney U *p*-value < 0.05 and Cliff’s delta ± 0.5 effect size). Plots organised by OPLS-DA modelling of females **A)** acute phase compared to non-burn controls (age 1) and **C)** ‘recovered’ compared to non-burn controls (age 3), with plots replicated for males in **B)** and **D)**. Grey lines show cut-off values for Cliff’s delta ± 0.5.

After recovery from the burn (>1 year post-injury), the lipidome of the burn injury cohort began to diverge when compared to non-burn controls at age 3. In the case of males, the OPLS-DA model initially failed to generate a significant orthogonal component and resulted in a lower Q^2^Y (cum) value of 0.108 when generation of an orthogonal component was forced (Table S2). Comparatively, the OPLS-DA model for the males at admission generated 3 orthogonal components while the models for females generated up to 5 orthogonal components (Table S2), which highlights the increased variation between female children post-injury compared to non-burn controls. This suggests that the lipidome of ‘recovered’ males was like that of non-burn controls, although some significant differences remained, particularly in the lysophosphatidylcholine (LPC) subclass. In contrast, females exhibited a distinct lipidomic signature at the ‘recovered’ stage, similar to our observations at the acute phase, with a robust model (R^2^X(cum) = 0.575, R^2^Y(cum) = 0.976, Q^2^Y(cum) = 0.527) and a clearer separation from non-burn controls.

Of particular interest was the role of the monoacylglycerol (MG) subclass which emerged as a key driver of separation in female paediatric burn patients at the ‘recovered’ stage. Over time, there was a marked decrease in MG species in females from the acute phase (adj. *p*-value < 0.001) to the ‘recovered’ stage, whereas this trend was not observed in males (Figure 2C-D, Figure S1). To investigate why the MG subclass had decreased in females, analysis shifted focus to the lipid species instead. Within the MG subclass, lipid species MG(18:1)(adj. *p*-value < 0.0001), MG(18:2) (adj. *p*-value < 0.0001) and MG(18:3) (adj. *p*-value < 0.01) showed the most significant decline over time for females, with levels lower than that of non-burn controls when ‘recovered’ (Figure S1). While males exhibited a similar pattern to females in the acute phase for the same MG species (adj. *p*-value < 0.0001), their levels recovered post-injury to match those of male non-burn controls at age 3 (adj. *p*-value > 0.05).

### Systemic inflammation is greater in females after a non-severe burn injury

As FAs were among the lipid subclasses significantly elevated in paediatric burn patients across both sexes during the acute phase and remained higher into recovery (Figure 2), we explored the specific species within this subclass given their roles in inflammation^52^. All 20 FA lipid species contributed substantially to group separation in the OPLS-DA models (Table S2), but of particular interest was arachidonic acid (FA(20:4)), which is a central precursor to an array of pro-inflammatory mediators^52^.

Longitudinal visualisation of FA(20:4) levels revealed sex-specific patterns (Figure 3A). In the acute phase, male and female burn patients exhibited significantly higher plasma concentrations of FA(20:4) compared to non-burn controls (adj. *p*-value < 0.0001). However, trajectories over time diverged for the sexes. In males, although FA(20:4) remained significantly elevated compared to non-burn male patients at age 3, we observed a gradual decline that appeared to be approaching the levels seen in non-burn controls. In contrast, females showed a persistent increase in FA(20:4) levels more than a year post-injury (‘recovered’), which is in contrast to the natural age-related decline of this FA that is observed in non-burn controls. This is indicative of a sex-specific disruption in lipid species following burn injury.

**Figure 3.**
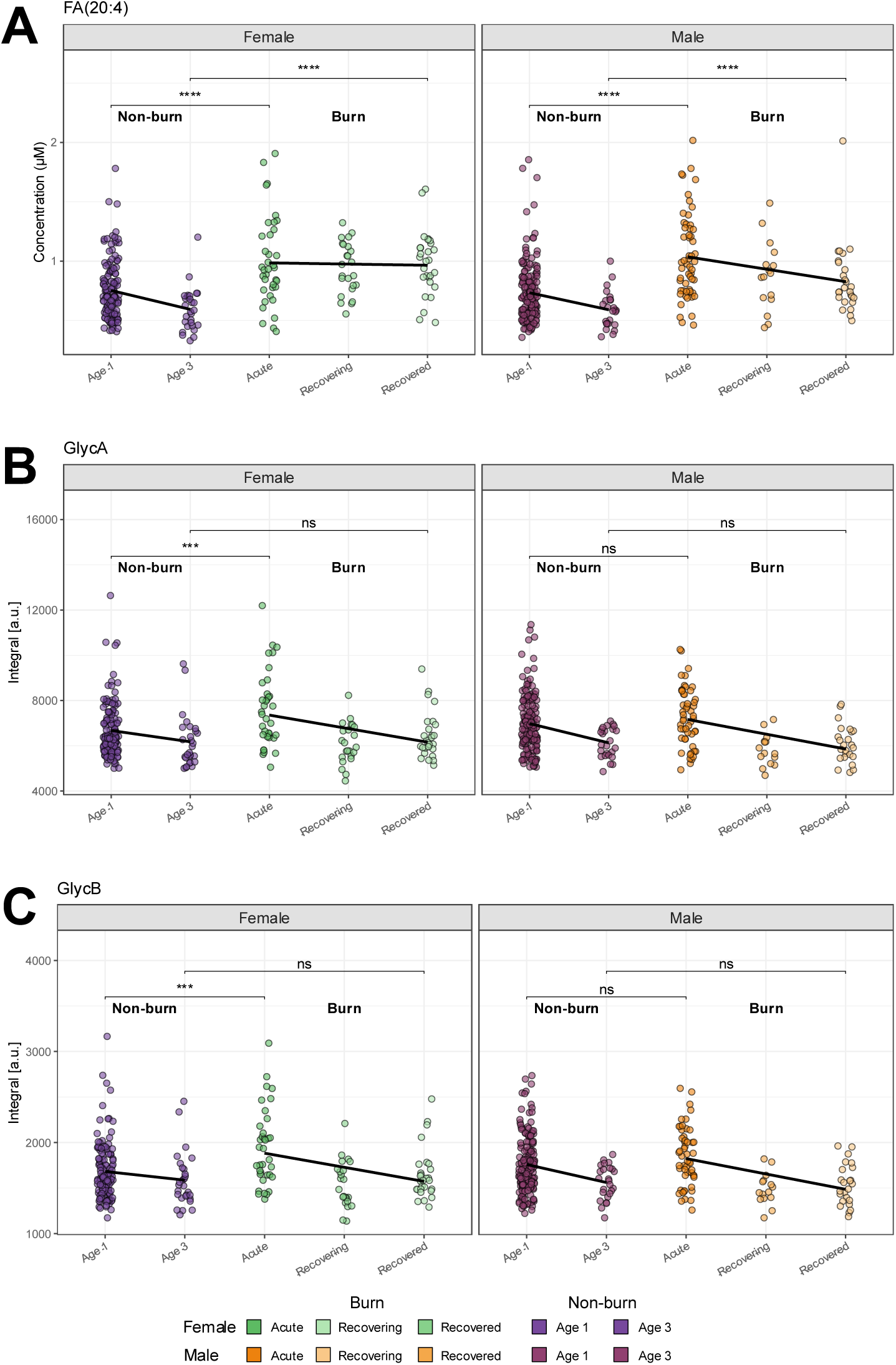
Sex differences in systemic inflammatory markers for paediatric burn injury in females and males compared to non-burn controls. Changes in females (green) compared to non-burn controls, replicated in males (orange) across the burn stages using a linear regression in **A)** FA(20:4), **B)** GlycA and **C)** GlycB. Univariate statistics using Mann-Whitney U (adjusted with Benjamin-Hochberg) show significance: ns = not significant, * = *p*-value < 0.05; ** = *p*-value < 0.01; *** = *p*-value < 0.001; **** = *p*-value < 0.0001.

Building on these findings, we assessed systemic inflammation through quantification of GlycA and GlycB, which are composite markers derived from *N*-glycan moieties that reflect circulating acute-phase glycoproteins during inflammation^53–55^. In the acute phase following burn injury, females showed significantly higher levels of both GlycA and GlycB (adj. *p*-value < 0.001) compared to female non-burn controls of a similar age (Figure 3B-C). GlycA and GlycB levels decreased over time in the recovering and recovered stages for females, eventually aligning with the levels in non-burn children. In contrast, male burn survivors showed no significant change in trajectory over time, with no significant increase in GlycA and GlycB in the acute phase and levels appearing to follow the typical developmental progression observed in non-burn male children aged between 1 and 3 years old. It has been demonstrated in healthy adults that there is no significant difference of the glycoproteins in relation to sex^56^.

### Long-term stress levels and lipid influence in response to paediatric burn injury

Cortisol measurements from hair samples were used as a non-invasive method to monitor long-term stress levels and to capture pre-injury baseline levels. Average cortisol levels in females and males were comparable from pre-injury (hair sample taken up to 1 week after injury) and remained so up to 3-6 months post-injury. Although the difference between males and females was not statistically significant, females demonstrated increasing variability over time in cortisol concentration as seen in the broader spread of their data up to 12+ months. This is reflected in the increasing fold changes from 1 week post-injury for females (3-6 months = 0.62 FC, 6-12 months = 2.27 FC and >12 months = 5.64 FC) compared to males who exhibited smaller fold changes in cortisol concentrations (3-6 months = 0.27 FC, 6-12 months = 0.83 FC and >12 months = 1.46 FC).

Correlation analysis of cortisol with 545 lipid species detected in their hair samples revealed sex-specific differences. In males, no significant associations were observed with only weak negative correlations across all lipid species (Figure 4B, Table S3), a trend also evident at the subclass level (Table S4). However, in females 101 lipid species were significantly correlated with cortisol after adjustment and 96% of which belonged to the TG subclass (Table S3, Table S4). The strongest association was observed for TG(56:2_FA20:0) (Pearson’s r = 0.72, adj. *p*-value = 1.77×10^−10^). The summed TG class also correlated significantly with cortisol (r = 0.38, adj. *p*-value = 0.015). Outside the TG subclass, only cholesteryl ester CE(18:2) showed a moderate association (r = 0.40, adj. *p*-value = 0.005).

**Figure 4.**
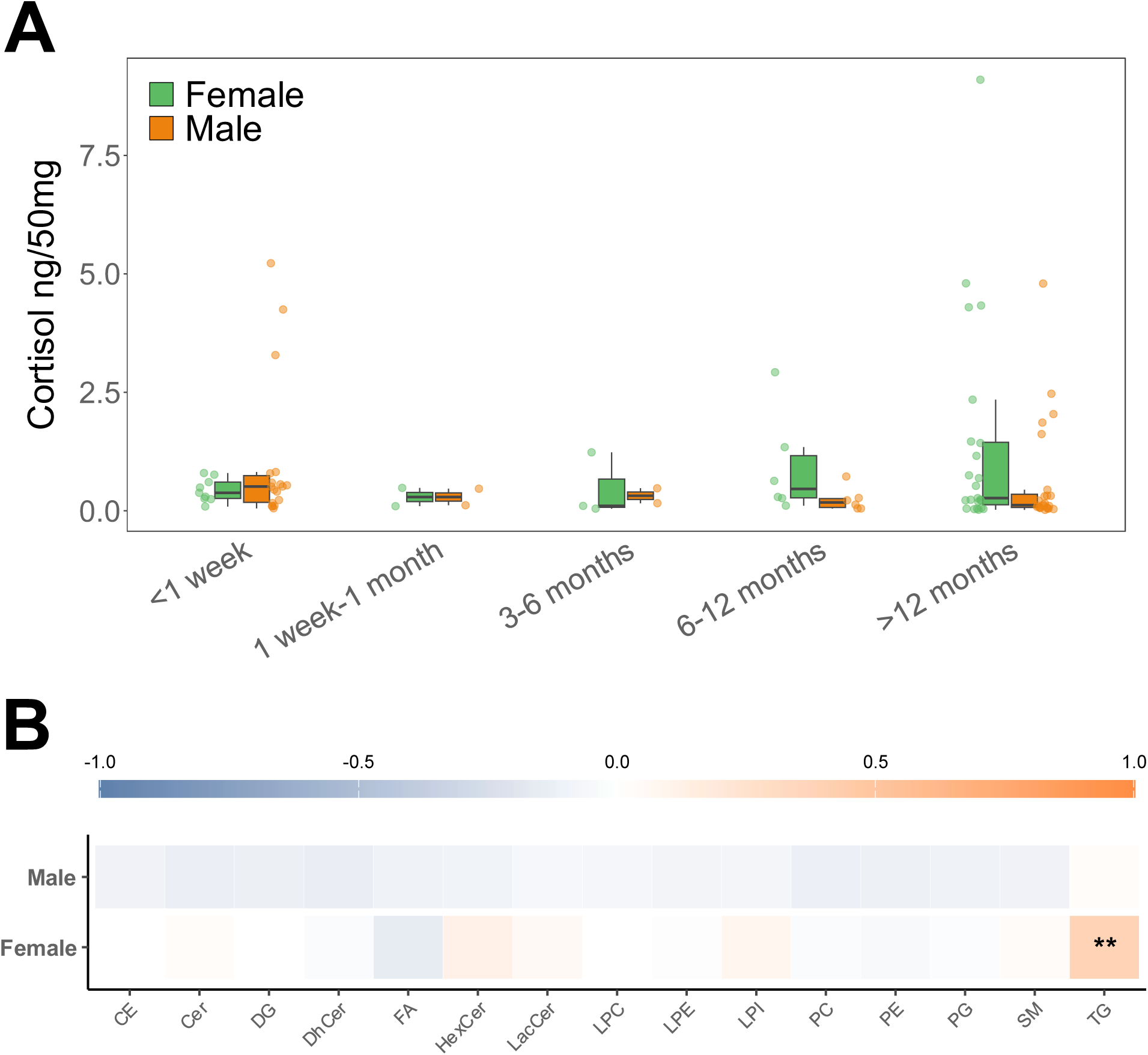
Cortisol trends and lipid subclass correlations split by sex in hair collected from the paediatric burn cohort. **A)** Females (green, n = 28) and males (orange, n = 35) cortisol measurements from 3 cm strands categorised into five timepoints (up to 1 week: n = 47; 1 week-1 month: n = 10; 3-6 months: n = 12; 6-12 months: n = 39; 12+ months: n = 42). Individual measurements for both sexes are indicated by the jittered dot points. **B)** Correlation matrix of lipid subclasses detected and cortisol levels in hair split by male and female children after burn injury. Pearson’s correlation scale ranging from positive (orange) to negative (blue) correlation with ‘*’ denoting a significant correlation after *p*-value adjustment for each lipid subclass.

## Discussion

This study highlights sex-specific trajectories of inflammation and lipid metabolism in paediatric burn patients. Consistent with prior research, we observed a disruption in the lipidome in both male and female children in the acute phase (<14 days) following burn injury, which was characterised by increased FA subclasses and a decrease in lysophospholipids (LPGs, LPCs and lysophosphatidylethanolamines (LPEs)) and phosphatidylglycerols (PGs). The shift in FAs is similar to lipidomic changes previously reported in adult^57^ and paediatric^58^ burn cohorts, which could indicate that perturbations in lipid metabolism, particularly increases in circulating FAs, are a hallmark of the early post-burn response regardless of age.

However, our data also revealed distinct sex differences in the paediatric burn plasma lipidome. Although both sexes showed lipidomic disruptions, males demonstrated a more pronounced increase in FA subclasses while females exhibited higher levels of the inflammatory glycoproteins GlycA and GlycB during the acute phase of the burn injury compared to non-burn controls. The elevated GlycA and GlycB levels in females that returned to non-burn control levels during recovery are consistent with a transient, systemic inflammatory response typically observed after a burn. These markers have been previously found by our research team to be increased post-burn in adults^59^ and children^6^. To our knowledge, no other studies have examined GlycA and GlycB responses specifically in paediatric burn trauma. Beyond burn injury, GlycA and GlycB have been reported to be elevated in adolescents with inflammatory conditions such as pre-diabetes^60^, obesity^61^ and cystic fibrosis^62^. Within these conditions, sex differences have also been observed in other traditional inflammatory markers, including C-reactive protein (CRP). In both adults and children, females consistently exhibited higher CRP levels than males during inflammatory responses which supports the sex-specific patterns observed in our study^27,63^. In contrast, males did not exhibit any spike in GlycA and GlycB during the acute phase or up to the ‘recovered’ stage. The implication of this muted systemic response is unknown, and it is unclear whether it predisposes males to a long-term recovery advantage or instead reflects an impeded activation of specific inflammatory pathways. Evidence for such observations is limited but findings from a study of post-operative systemic inflammation following major abdominal surgery showed that higher systemic inflammation (measured by CRP) was associated with poorer quality of recovery for adults^64^. This raises the possibility that the minimal systemic response observed in male children may be more protective in the context of burn recovery, but more research is needed in this area.

The increases in FA lipid species observed in children with burn injury may also indicate a shift to a pro-inflammatory state. Within the FA subclasses, there was a lack of elevation in some omega-3 fatty acids, particularly eicosapentaenoic acid (FA(20:5)) and (FA(22:6)), during both the acute and recovery stages. These long-chain polyunsaturated fatty acids are known precursors of anti-inflammatory mediators, such as resolvins and protectins, that modulate inflammation^65^. This is coupled with an increase in FA(20:4), also known as arachidonic acid, which is a precursor to numerous pro-inflammatory molecules in the eicosanoid pathway, including prostaglandins, thromboxanes, and leukotrienes^65^. The skewed ratio with more pro-inflammatory FA(20:4) to anti-inflammatory FA(20:5) or FA(22:6) suggests that the paediatric burn lipidome remains in a pro-inflammatory state even when considered ‘recovered’. Notably, although still elevated, FA(20:4) levels in males appear to trend toward non-burn control levels one year post-injury. In contrast, females had a persistent, unchanged increase in FA(20:4) even during the ‘recovered’ stage.

Additionally, females had a unique lipidomic change compared to non-burn controls that males did not exhibit when considered ‘recovered’. One-year post-burn, we observed a significant depletion of the MG subclasses in female burn survivors, which was not observed in males. In contrast, our team has previously identified MG lipid species in adult burn patients to increase after injury^59^ which demonstrates an opposing response in paediatric female burn survivors. This decrease in the MG subclass and increased FAs, particularly 1-MG species (MG(18:1), MG(18:2) and MG(18:3)), may suggest a sex-specific disruption in the *de novo* synthesis of MGs, potentially via impaired esterification of FAs to glycerol^66^ and could reflect a maladaptive lipidomic response during burn recovery.

In this study, we have identified sex-specific lipidomic changes in paediatric burns that may be influenced inherently by sex hormones^13^. Sex differences in burn recovery outcomes have been attributed to the role of oestrogen and related hormones as potent modulators of inflammation^67–69^. However, limited studies conducted mainly in murine models implies the biological mechanisms of these sex-based disparities are not well understood, particularly in pre-pubescent children where the role of oestrogen and other sex hormones is less clear. We propose that hormones are not the sole drivers of sex differences in paediatric burn outcomes, as sex-dependent hormonal activity is unlikely to play a major role in children at such a young age. Blood oestradiol and testosterone levels typically rise significantly only around age 8 in females and age 9 in males, respectively^70^. Thus, other factors may substantially contribute to these sex-specific differences. One important consideration is stress which represents a major long-term health issue following both severe and non-severe burn injuries in both adults and children^71–73^. When comparing cortisol hair levels over time in the paediatric burn cohort, females displayed a trend of increased cortisol levels and greater inter-individual variability from 6-12 months to over a year post-injury. Females who experienced a burn in their childhood have been reported to be twice as likely than males to develop a psychological disorder, such as an anxiety disorder, post-traumatic stress disorder or depression^16,74^. This trend may indicate a prolonged stress response in females which aligns with prior findings that female paediatric burn survivors report lower quality of life^75^.

Moreover, this trend may be linked to persistent elevations in arachidonic acid (FA 20:4), a precursor to eicosanoids, which can mediate stress and inflammation^76^. On the contrary, hair cortisol levels did not increase in males from pre-injury (hair sampled up to a week post-injury) to a year post-burn. There are some discrepancies in the literature where studies have identified higher levels of urinary cortisol in males after a burn than females^77,78^. It is noted that hair cortisol levels do not associate with urinary cortisol but are more closely aligned to salivary cortisol, which may explain the differing results in the literature regarding cortisol measurements from hair, saliva or urine post-injury^79^. Interestingly, we identified a potential correlation between hair lipids and cortisol measurements only in female children post-burn injury. TG species showed the most significant and strongest correlations in this group that potentially aligns with limited evidence in other inflammatory conditions such as paediatric obesity, where circulating triglycerides positively associate with cortisol levels^80^. In adults, stressors can stimulate the HPA axis and lead to elevated cortisol levels that in turn raise circulating triglycerides^81^. These findings suggest a possible link between circulating TGs and cortisol that can be detected in hair which can offer a novel approach for monitoring stress and systemic lipid changes. Moreover, the observed associations highlight a sex-specific relationship between stress and inflammation that warrants further investigation, as males showed no significant correlations between cortisol and lipids that is likely reflected by their relatively stable cortisol levels.

Importantly, our findings contribute to a broader and sometimes contradictory discussion on sex-specific outcomes in burn recovery. Some studies have reported that females fare worse post-burn in terms of physical and mental health, psychological distress, and burn-specific quality of life metrics^82–84^. Other studies suggest that females exhibit a less profound hypermetabolic response compared to males through reduced catecholamines, energy expenditure, and protein catabolism which could be protective^78^. The dichotomy of these perspectives is complex and underpins the necessity for validating these findings in population-based paediatric burn cohorts to fully elucidate the potential differences between sexes in response to injury.

The role of demographic and injury-related factors must also be considered. Previous studies have linked female sex, young age, and specific burn sites (e.g., face, neck and upper limbs) with increased risk of poor long-term outcomes, including pathological scarring^14^. In this cohort, females experienced significantly higher face and neck burns than males. These factors could influence the lipidomic and inflammatory alterations we observed, potentially related to the psychological stress of having visible burns but this topic remains a subject of debate in the literature^73,85^. Our findings also emphasise that mental health should not be overlooked in young children post-burn, as psychological and physiological stress responses may shape recovery in ways that are not immediately evident in clinical assessments. Notably, we observed sex-specific differences in lipidomic profiles and cortisol levels in response to relatively minor (non-severe) burns, as the children had an average of 2% TBSA. This suggests that even non-severe burn injuries can trigger systemic responses in children as previously seen with potential long-term psychological and physiological consequences^6,7,38,73,86,87^. These findings warrant greater attention to childhood non-severe burn injury in both research and clinical settings. We have identified unique, sex-specific lipidomic and inflammatory trajectories, along with stress-related trends that may deepen our understanding of the metabolic consequences of post-paediatric non-severe burn injury. Further studies should link these changes to long-term outcomes and clarify how sex, stress, and inflammation connect during paediatric burn recovery.

## Conclusion

This study identified sex-specific differences in lipidomic and inflammatory responses following non-severe burn injury in paediatric patients. While both males and females showed disruptions in lipid metabolism during the acute phase, only females exhibited persistent alterations with elevated FA(20:4) (arachidonic acid) and depleted MG lipid species over one year post-injury, despite their burn being fully healed. These changes were accompanied by higher levels of systemic inflammatory markers GlycA and GlycB during the acute phase in females and a potential trend toward elevated long-term stress through hair cortisol levels that significantly correlation with TG species. In contrast, males showed a more transient inflammatory profile, with lipid levels becoming similar to those of non-burn controls in the ‘recovered’ stage, with the exception of change in plasma FA concentrations, which was trending downward. Although clinical characteristics between sexes were similar, the observed biological differences suggest that sex may play a role in shaping long-term recovery trajectories at the molecular level. Burn trauma and the impact of factors like stress and injury location on recovery is complex and our findings underscore the need for population-based, longitudinal studies to validate these sex-specific patterns and explore their implications for outcomes and therapeutic strategies in paediatric care with non-severe burns.

## Data Availability

The data presented in this study are not publicly available. Data are available from ORIGINS via an application process, including Scientific Committee and ORIGINS Director approval, Ramsay Human Research Ethics approval, and payment of an access fee. This application process is in place as the data collected are sensitive health data related to pregnancy, birth and children.

## Acknowledgements and Funding

We dedicate this work to the memory of our co-author and mentor, Dr. Mark W Fear. His guidance, dedication and commitment to pushing the boundaries of research and science for the betterment of children’s health has left an imprint on all who worked with him. His passion for improving the lives of children and families continues to inspire us and his legacy will forever guide our work.

We are grateful to all the ORIGINS families who support the project. We would also like to acknowledge and thank the following teams and individuals who have made ORIGINS possible: ORIGINS project team; Joondalup Health Campus (JHC); members of ORIGINS Community Reference and Participant Reference Groups; Research Interest Groups and the ORIGINS Scientific Committee; The Kids Research Institute Australia; City of Wanneroo; City of Joondalup; and Professor Fiona Stanley. This study is a sub-project of ORIGINS. This unique long-term study, a collaboration between The Kids Research Institute Australia and Joondalup Health Campus, is one of the most comprehensive studies of pregnant women and their families in Australia to date, recruiting 10,000 families over a decade from the Joondalup and Wanneroo communities of Western Australia.

ORIGINS has received core funding support from the Telethon Perth Children’s Hospital Research Fund, Joondalup Health Campus, the Paul Ramsay Foundation and the Commonwealth Government of Australia through the Channel 7 Telethon Trust. Substantial in-kind support has been provided by The Kids Research Institute Australia and Joondalup Health Campus. Additionally, the Children’s Burn Injury Biobank has received core funding support from the Fiona Wood Foundation and the Stan Perron Charitable Foundation. Sample preparation and mass spectrometry analyses conducted at the Australian National Phenome Centre were supported by the Western Australian State Government and the Medical Research Future Fund. Author Holmes acknowledges support from the Department of Jobs, Tourism, Science and Innovation through the Western Australian Premier’s Fellowship, as well as the Australian Research Council Laureate Fellowship. Author Whiley acknowledges support from Dementia Australia and the Royce Simmons Foundation through the mid-career fellowship.

## References

(1) Duke, J. M.; Rea, S.; Boyd, J. H.; Randall, S. M.; Wood, F. M. Mortality After Burn Injury in Children: A 33-Year Population-Based Study. Pediatrics 2015, 135 (4), e903–e910. 10.1542/peds.2014-3140.

(2) Duke, J. M.; Randall, S. M.; Vetrichevvel, T. P.; McGarry, S.; Boyd, J. H.; Rea, S.; Wood, F. M. Long-Term Mental Health Outcomes after Unintentional Burns Sustained during Childhood: A Retrospective Cohort Study. Burns Trauma 2018, 6, 32. 10.1186/s41038-018-0134-z.

(3) Duke, J. M.; Randall, S. M.; Fear, M. W.; Boyd, J. H.; Rea, S.; Wood, F. M. Diabetes Mellitus after Injury in Burn and Non-Burned Patients: A Population Based Retrospective Cohort Study. Burns 2018, 44 (3), 566–572. 10.1016/j.burns.2017.10.019.

(4) Hundeshagen, G.; Herndon, D. N.; Clayton, R. P.; Wurzer, P.; McQuitty, A.; Jennings, K.; Branski, L.; Collins, V. N.; Marques, N. R.; Finnerty, C. C.; Suman, O. E.; Kinsky, M. P. Long-Term Effect of Critical Illness after Severe Paediatric Burn Injury on Cardiac Function in Adolescent Survivors: An Observational Study. Lancet Child Adolesc Health 2017, 1 (4), 293–301. 10.1016/S2352-4642(17)30122-0.

(5) Barrett, L. W.; Fear, V. S.; Waithman, J. C.; Wood, F. M.; Fear, M. W. Understanding Acute Burn Injury as a Chronic Disease. Burns & Trauma 2019, 7, s41038-019-0163–2. 10.1186/s41038-019-0163-2.

(6) Begum, S.; Lodge, S.; Hall, D.; Johnson, B. Z.; Bong, S. H.; Whiley, L.; Gray, N.; Fear, V. S.; Fear, M. W.; Holmes, E.; Wood, F. M.; Nicholson, J. K. Cardiometabolic Disease Risk Markers Are Increased Following Burn Injury in Children. Front Public Health 2023, 11, 1105163. 10.3389/fpubh.2023.1105163.

(7) Johnson, B. Z.; McAlister, S.; McGuire, H. M.; Palanivelu, V.; Stevenson, A.; Richmond, P.; Palmer, D. J.; Metcalfe, J.; Prescott, S. L.; Wood, F. M.; Fazekas de St Groth, B.; Linden, M. D.; Fear, M. W.; Fear, V. S. Pediatric Burn Survivors Have Long-Term Immune Dysfunction With Diminished Vaccine Response. Front. Immunol. 2020, 11. 10.3389/fimmu.2020.01481.

(8) McGwin, G.; George, R. L.; Cross, J. M.; Reiff, D. A.; Chaudry, I. H.; Rue, L. W. Gender Differences in Mortality Following Burn Injury. Shock 2002, 18 (4), 311–315. 10.1097/00024382-200210000-00004.

(9) Mehta, K.; Arega, H.; Smith, N. L.; Li, K.; Gause, E.; Lee, J.; Stewart, B. Gender-Based Disparities in Burn Injuries, Care and Outcomes: A World Health Organization (WHO) Global Burn Registry Cohort Study. Am J Surg 2022, 223 (1), 157–163. 10.1016/j.amjsurg.2021.07.041.

(10) Kumar, P.; Chirayil, P. T.; Chittoria, R. Ten Years Epidemiological Study of Paediatric Burns in Manipal, India. Burns 2000, 26 (3), 261–264. 10.1016/s0305-4179(99)00109-6.

(11) Moore, E. C.; Pilcher, D.; Bailey, M.; Cleland, H. Women Are More than Twice as Likely to Die from Burns as Men in Australia and New Zealand: An Unexpected Finding of the Burns Evaluation And Mortality (BEAM) Study. J Crit Care 2014, 29 (4), 594–598. 10.1016/j.jcrc.2014.03.021.

(12) Tedesco, D. J.; Hutter, M. F.; Khalaf, F.; Pond, G. R.; Jeschke, M. G. Sex- and Age-Related Differences in Post-Burn Pathophysiology. Critical Care Medicine 10.1097/CCM.0000000000006789. 10.1097/CCM.0000000000006789.

(13) Summers, J. I.; Ziembicki, J. A.; Corcos, A. C.; Peitzman, A. B.; Billiar, T. R.; Sperry, J. L. Characterization of Sex Dimorphism Following Severe Thermal Injury. J Burn Care Res 2014, 35 (6), 484–490. 10.1097/BCR.0000000000000018.

(14) Gangemi, E. N.; Gregori, D.; Berchialla, P.; Zingarelli, E.; Cairo, M.; Bollero, D.; Ganem, J.; Capocelli, R.; Cuccuru, F.; Cassano, P.; Risso, D.; Stella, M. Epidemiology and Risk Factors for Pathologic Scarring after Burn Wounds. Arch Facial Plast Surg 2008, 10 (2), 93–102. 10.1001/archfaci.10.2.93.

(15) Foley, P.; Jeeves, A.; Davey, R. B.; Sparnon, A. L. Breast Burns Are Not Benign: Long-Term Outcomes of Burns to the Breast in Pre-Pubertal Girls. Burns 2008, 34 (3), 412–417. 10.1016/j.burns.2007.05.001.

(16) Goodhew, F.; Van Hooff, M.; Sparnon, A.; Roberts, R.; Baur, J.; Saccone, E. J.; McFarlane, A. Psychiatric Outcomes amongst Adult Survivors of Childhood Burns. Burns 2014, 40 (6), 1079–1088. 10.1016/j.burns.2014.04.017.

(17) Pope, S. J.; Solomons, W. R.; Done, D. J.; Cohn, N.; Possamai, A. M. Body Image, Mood and Quality of Life in Young Burn Survivors. Burns 2007, 33 (6), 747–755. 10.1016/j.burns.2006.10.387.

(18) Jeschke, M. G.; Mlcak, R. P.; Finnerty, C. C.; Norbury, W. B.; Przkora, R.; Kulp, G. A.; Gauglitz, G. G.; Zhang, X.-J.; Herndon, D. N. Gender Differences in Pediatric Burn Patients. Ann Surg 2008, 248 (1), 126–136. 10.1097/SLA.0b013e318176c4b3.

(19) Hussain, A.; Dunn, K. Burn Related Mortality in Greater Manchester: 11-Year Review of Regional Coronial Department Data. Burns 2015, 41 (2), 225–234. 10.1016/j.burns.2014.10.008.

(20) Phelan, H. A.; Shafi, S.; Parks, J.; Maxson, R. T.; Ahmad, N.; Murphy, J. T.; Minei, J. P. Use of a Pediatric Cohort to Examine Gender and Sex Hormone Influences on Outcome After Trauma. Journal of Trauma and Acute Care Surgery 2007, 63 (5), 1127. 10.1097/TA.0b013e318154c1b8.

(21) Jeschke, M. G.; Przkora, R.; Suman, O. E.; Finnerty, C. C.; Mlcak, R. P.; Pereira, C. T.; Sanford, A. P.; Herndon, D. N. Sex differences in the long-term outcome after a severe thermal injury. Shock 2007, 27 (5), 461. 10.1097/01.shk.0000238071.74524.9a.

(22) Bernardi, S.; Marcuzzi, A.; Piscianz, E.; Tommasini, A.; Fabris, B. The Complex Interplay between Lipids, Immune System and Interleukins in Cardio-Metabolic Diseases. Int J Mol Sci 2018, 19 (12), 4058. 10.3390/ijms19124058.

(23) Hao, Y.; Li, D.; Xu, Y.; Ouyang, J.; Wang, Y.; Zhang, Y.; Li, B.; Xie, L.; Qin, G. Investigation of Lipid Metabolism Dysregulation and the Effects on Immune Microenvironments in Pan-Cancer Using Multiple Omics Data. BMC Bioinformatics 2019, 20 (Suppl 7), 195. 10.1186/s12859-019-2734-4.

(24) Dong, J.; Yang, S.; Zhuang, Q.; Sun, J.; Wei, P.; Zhao, X.; Chen, Y.; Chen, X.; Li, M.; Wei, L.; Chen, C.; Fan, Y.; Shen, C. The Associations of Lipid Profiles With Cardiovascular Diseases and Death in a 10-Year Prospective Cohort Study. Front Cardiovasc Med 2021, 8, 745539. 10.3389/fcvm.2021.745539.

(25) Erion, D. M.; Park, H.-J.; Lee, H.-Y. The Role of Lipids in the Pathogenesis and Treatment of Type 2 Diabetes and Associated Co-Morbidities. BMB Rep 2016, 49 (3), 139–148. 10.5483/BMBRep.2016.49.3.268.

(26) Leung, B.; Younger, J. F.; Stockton, K.; Muller, M.; Paratz, J. Cardiovascular Risk Profile in Burn Survivors. Burns 2017, 43 (7), 1411–1417. 10.1016/j.burns.2017.07.010.

(27) Casimir, G. J. A.; Mulier, S.; Hanssens, L.; Zylberberg, K.; Duchateau, J. Gender differences in inflammatory markers in children. Shock 2010, 33 (3), 258. 10.1097/SHK.0b013e3181b2b36b.

(28) Casimir, G. J. A.; Mulier, S.; Hanssens, L.; Knoop, C.; Ferster, A.; Hofman, B.; Duchateau, J. Chronic inflammatory diseases in children are more severe in girls. Shock 2010, 34 (1), 23. 10.1097/SHK.0b013e3181ce2c3d.

(29) LaBarre, J. L.; Miller, A. L.; Bauer, K. W.; Burant, C. F.; Lumeng, J. C. Early Life Stress Exposure Associated with Reduced Polyunsaturated-Containing Lipids in Low-Income Children. Pediatr Res 2021, 89 (5), 1310–1315. 10.1038/s41390-020-0989-0.

(30) Davis, S. L.; Latimer, M.; Rice, M. Biomarkers of Stress and Inflammation in Children. Biol Res Nurs 2023, 25 (4), 559–570. 10.1177/10998004231168805.

(31) Tsao, J. C. I.; Evans, S.; Meldrum, M.; Zeltzer, L. K. Sex Differences in Anxiety Sensitivity among Children with Chronic Pain and Non-Clinical Children. J Pain Manag 2009, 2 (2), 151–161. Retrieved from https://pmc.ncbi.nlm.nih.gov/articles/PMC3072583/

(32) Noll, J. G.; Zeller, M. H.; Trickett, P. K.; Putnam, F. W. Obesity Risk for Female Victims of Childhood Sexual Abuse: A Prospective Study. Pediatrics 2007, 120 (1), e61–e67. 10.1542/peds.2006-3058.

(33) Pervanidou, P.; Chrousos, G. P. Metabolic Consequences of Stress during Childhood and Adolescence. Metabolism 2012, 61 (5), 611–619. 10.1016/j.metabol.2011.10.005.

(34) Liu, J.; Dai, Y.; Yuan, E.; Li, Y.; Wang, Q.; Wang, L.; Su, Y. Age-Specific and Sex-Specific Reference Intervals for Non-Fasting Lipids and Apolipoproteins in 7260 Healthy Chinese Children and Adolescents Measured with an Olympus AU5400 Analyser: A Cross-Sectional Study. BMJ Open 2019, 9 (8), e030201. 10.1136/bmjopen-2019-030201.

(35) Holven, K. B.; Roeters van Lennep, J. Sex Differences in Lipids: A Life Course Approach. Atherosclerosis 2023, 384, 117270. 10.1016/j.atherosclerosis.2023.117270.

(36) Tabassum, R.; Ruotsalainen, S.; Ottensmann, L.; Gerl, M. J.; Klose, C.; Tukiainen, T.; Pirinen, M.; Simons, K.; Widén, E.; Ripatti, S. Lipidome- and Genome-Wide Study to Understand Sex Differences in Circulatory Lipids. Journal of the American Heart Association 2022, 11 (19), e027103. 10.1161/JAHA.122.027103.

(37) Medina, J.; Goss, N.; Correia, G. dos S.; Borreggine, R.; Teav, T.; Kutalik, Z.; Vidal, P. M.; Gallart-Ayala, H.; Ivanisevic, J. Clinical Lipidomics Reveals High Individuality and Sex Specificity of Circulatory Lipid Signatures: A Prospective Healthy Population Study. Journal of Lipid Research 2025, 66 (5), 100780. 10.1016/j.jlr.2025.100780.

(38) Kierath, E.; Ryan, M.; Holmes, E.; Nicholson, J. K.; Fear, M. W.; Wood, F. M.; Whiley, L.; Gray, N. Plasma Lipidomics Reveal Systemic Changes Persistent throughout Early Life Following a Childhood Burn Injury. BURNS TRAUMA 2023, 11, tkad044. 10.1093/burnst/tkad044.

(39) Silva, D. T.; Hagemann, E.; Davis, J. A.; Gibson, L. Y.; Srinivasjois, R.; Palmer, D. J.; Colvin, L.; Tan, J.; Prescott, S. L. Introducing the ORIGINS Project: A Community-Based Interventional Birth Cohort. Reviews on Environmental Health 2020, 35 (3), 281–293. 10.1515/reveh-2020-0057.

(40) D’Vaz, N.; Kidd, C.; Miller, S.; Amin, M.; Davis, J. A.; Talati, Z.; Silva, D. T.; Prescott, S. L. The ORIGINS Project Biobank: A Collaborative Bio Resource for Investigating the Developmental Origins of Health and Disease. International Journal of Environmental Research and Public Health 2023, 20 (13), 6297. 10.3390/ijerph20136297.

(41) Ryan, M. J.; Grant-St James, A.; Lawler, N. G.; Fear, M. W.; Raby, E.; Wood, F. M.; Maker, G. L.; Wist, J.; Holmes, E.; Nicholson, J. K.; Whiley, L.; Gray, N. Comprehensive Lipidomic Workflow for Multicohort Population Phenotyping Using Stable Isotope Dilution Targeted Liquid Chromatography-Mass Spectrometry. Journal of Proteome Research 2023, 22 (5), 1419–1433. 10.1021/acs.jproteome.2c00682.

(42) R Core Team. R: A Language and Environment for Statistical Computing., 2024. https://www.R-project.org/.

(43) Dona, A. C.; Jiménez, B.; Schäfer, H.; Humpfer, E.; Spraul, M.; Lewis, M. R.; Pearce, J. T. M.; Holmes, E.; Lindon, J. C.; Nicholson, J. K. Precision High-Throughput Proton NMR Spectroscopy of Human Urine, Serum, and Plasma for Large-Scale Metabolic Phenotyping. Anal. Chem. 2014, 86 (19), 9887–9894. 10.1021/ac5025039.

(44) Lodge, S.; Nitschke, P.; Kimhofer, T.; Wist, J.; Bong, S.-H.; Loo, R. L.; Masuda, R.; Begum, S.; Richards, T.; Lindon, J. C.; Bermel, W.; Reinsperger, T.; Schaefer, H.; Spraul, M.; Holmes, E.; Nicholson, J. K. Diffusion and Relaxation Edited Proton NMR Spectroscopy of Plasma Reveals a High-Fidelity Supramolecular Biomarker Signature of SARS-CoV-2 Infection. Anal. Chem. 2021, 93 (8), 3976–3986. 10.1021/acs.analchem.0c04952.

(45) Liu, M.; Nicholson, J. K.; Lindon, J. C. High-Resolution Diffusion and Relaxation Edited One- and Two-Dimensional 1H NMR Spectroscopy of Biological Fluids. Anal Chem 1996, 68 (19), 3370–3376. 10.1021/ac960426p.

(46) Masuda, R.; Lodge, S.; Whiley, L.; Gray, N.; Lawler, N.; Nitschke, P.; Bong, S.-H.; Kimhofer, T.; Loo, R. L.; Boughton, B.; Zeng, A. X.; Hall, D.; Schaefer, H.; Spraul, M.; Dwivedi, G.; Yeap, B. B.; Diercks, T.; Bernardo-Seisdedos, G.; Mato, J. M.; Lindon, J. C.; Holmes, E.; Millet, O.; Wist, J.; Nicholson, J. K. Exploration of Human Serum Lipoprotein Supramolecular Phospholipids Using Statistical Heterospectroscopy in N-Dimensions (SHY-n): Identification of Potential Cardiovascular Risk Biomarkers Related to SARS-CoV-2 Infection. Analytical Chemistry 2022, 94 (10), 4426–4436. 10.1021/acs.analchem.1c05389.

(47) Bell, J. D.; Brown, J. C.; Nicholson, J. K.; Sadler, P. J. Assignment of Resonances for “acute-Phase” Glycoproteins in High Resolution Proton NMR Spectra of Human Blood Plasma. FEBS Lett 1987, 215 (2), 311–315. 10.1016/0014-5793(87)80168-0.

(48) Otvos, J. D.; Shalaurova, I.; Wolak-Dinsmore, J.; Connelly, M. A.; Mackey, R. H.; Stein, J. H.; Tracy, R. P. GlycA: A Composite Nuclear Magnetic Resonance Biomarker of Systemic Inflammation. Clin Chem 2015, 61 (5), 714–723. 10.1373/clinchem.2014.232918.

(49) Kimhofer, T.; Lodge, S.; Whiley, L.; Gray, N.; Loo, R. L.; Lawler, N. G.; Nitschke, P.; Bong, S. H.; Morrison, D. L.; Begum, S.; Richards, T.; Yeap, B. B.; Smith, C.; Smith, K. G. C.; Holmes, E.; Nicholson, J. K. Integrative Modeling of Quantitative Plasma Lipoprotein, Metabolic, and Amino Acid Data Reveals a Multiorgan Pathological Signature of SARS-CoV-2 Infection. Journal of Proteome Research 2020, 19 (11), 4442–4454. 10.1021/ACS.JPROTEOME.0C00519.

(50) Ghorasaini, M.; Mohammed, Y.; Adamski, J.; Bettcher, L.; Bowden, J. A.; Cabruja, M.; Contrepois, K.; Ellenberger, M.; Gajera, B.; Haid, M.; Hornburg, D.; Hunter, C.; Jones, C. M.; Klein, T.; Mayboroda, O.; Mirzaian, M.; Moaddel, R.; Ferrucci, L.; Lovett, J.; Nazir, K.; Pearson, M.; Ubhi, B. K.; Raftery, D.; Riols, F.; Sayers, R.; Sijbrands, E. J. G.; Snyder, M. P.; Su, B.; Velagapudi, V.; Williams, K. J.; de Rijke, Y. B.; Giera, M. Cross-Laboratory Standardization of Preclinical Lipidomics Using Differential Mobility Spectrometry and Multiple Reaction Monitoring. Anal. Chem. 2021, 93 (49), 16369–16378. 10.1021/acs.analchem.1c02826.

(51) Wood, F. M.; Phillips, M.; Jovic, T.; Cassidy, J. T.; Cameron, P.; Edgar, D. W.; Zealand (BRANZ), S. C. of the B. R. of A. and N. Water First Aid Is Beneficial In Humans Post-Burn: Evidence from a Bi-National Cohort Study. PLOS ONE 2016, 11 (1), e0147259. 10.1371/journal.pone.0147259.

(52) Wang, B.; Wu, L.; Chen, J.; Dong, L.; Chen, C.; Wen, Z.; Hu, J.; Fleming, I.; Wang, D. W. Metabolism Pathways of Arachidonic Acids: Mechanisms and Potential Therapeutic Targets. Sig Transduct Target Ther 2021, 6 (1), 1–30. 10.1038/s41392-020-00443-w.

(53) Nitschke, P.; Lodge, S.; Kimhofer, T.; Masuda, R.; Bong, S.-H.; Hall, D.; Schäfer, H.; Spraul, M.; Pompe, N.; Diercks, T.; Bernardo-Seisdedos, G.; Mato, J. M.; Millet, O.; Susic, D.; Henry, A.; El-Omar, E. M.; Holmes, E.; Lindon, J. C.; Nicholson, J. K.; Wist, J. J-Edited DIffusional Proton Nuclear Magnetic Resonance Spectroscopic Measurement of Glycoprotein and Supramolecular Phospholipid Biomarkers of Inflammation in Human Serum. Anal. Chem. 2022, 94 (2), 1333–1341. 10.1021/acs.analchem.1c04576.

(54) Fuertes-Martín, R.; Correig, X.; Vallvé, J.-C.; Amigó, N. Human Serum/Plasma Glycoprotein Analysis by 1H-NMR, an Emerging Method of Inflammatory Assessment. J Clin Med 2020, 9 (2), 354. 10.3390/jcm9020354.

(55) Lodge, S.; Masuda, R.; Nitschke, P.; Beilby, J. P.; Hui, J.; Hunter, M.; Litton, E.; Raby, E.; Currie, A.; Yeap, B. B.; Millet, O.; Cannet, C.; Schaefer, H.; Spraul, M.; Holmes, E.; Wist, J.; Nicholson, J. K. NMR Spectroscopy-Based Lipoprotein and Glycoprotein Biomarkers Differentiate Acute and Chronic Inflammation in Diverse Healthy and Disease Population Cohorts. J. Proteome Res. 2025, 24 (8), 4191–4201. 10.1021/acs.jproteome.5c00300.

(56) Lodge, S.; Masuda, R.; Nitschke, P.; Beilby, J. P.; Hui, J.; Hunter, M.; Yeap, B. B.; Millet, O.; Wist, J.; Nicholson, J. K.; Holmes, E. NMR Spectroscopy Derived Plasma Biomarkers of Inflammation in Human Populations: Influences of Age, Sex and Adiposity. PLOS ONE 2025, 20 (1), e0311975. 10.1371/journal.pone.0311975.

(57) Qi, P.; Abdullahi, A.; Stanojcic, M.; Patsouris, D.; Jeschke, M. G. Lipidomic Analysis Enables Prediction of Clinical Outcomes in Burn Patients. Sci Rep 2016, 6 (1), 38707. 10.1038/srep38707.

(58) Kraft, R.; Herndon, D. N.; Finnerty, C. C.; Hiyama, Y.; Jeschke, M. G. Association of Postburn Fatty Acids and Triglycerides with Clinical Outcome in Severely Burned Children. J Clin Endocrinol Metab 2013, 98 (1), 314–321. 10.1210/jc.2012-2599.

(59) Ryan, M. J.; Raby, E.; Whiley, L.; Masuda, R.; Lodge, S.; Nitschke, P.; Maker, G. L.; Wist, J.; Holmes, E.; Wood, F. M.; Nicholson, J. K.; Fear, M. W.; Gray, N. Nonsevere Burn Induces a Prolonged Systemic Metabolic Phenotype Indicative of a Persistent Inflammatory Response Postinjury. J. Proteome Res. 2024, 23 (8), 2893–2907. 10.1021/acs.jproteome.3c00516.

(60) Olson, M. L.; Rentería-Mexía, A.; Connelly, M. A.; Vega-López, S.; Soltero, E. G.; Konopken, Y. P.; Williams, A. N.; Castro, F. G.; Keller, C. S.; Yang, H. P.; Todd, M. W.; Shaibi, G. Q. Decreased GlycA Following Lifestyle Intervention Among Obese, Prediabetic Adolescent Latinos. J Clin Lipidol 2019, 13 (1), 186–193. 10.1016/j.jacl.2018.09.011.

(61) Jago, R.; Drews, K. L.; Otvos, J. D.; Willi, S. M.; Buse, J. B. Novel Measures of Inflammation and Insulin Resistance Are Related to Obesity and Fitness in a Diverse Sample of 11–14 Year-Olds: The HEALTHY Study. Int J Obes (Lond) 2016, 40 (7), 1157–1163. 10.1038/ijo.2016.84.

(62) Kevat, A. C.; Carzino, R.; Vidmar, S.; Ranganathan, S. Glycoprotein A as a Biomarker of Pulmonary Infection and Inflammation in Children with Cystic Fibrosis. Pediatric Pulmonology 2020, 55 (2), 401–406. 10.1002/ppul.24558.

(63) Khera, A.; Vega, G. L.; Das, S. R.; Ayers, C.; McGuire, D. K.; Grundy, S. M.; de Lemos, J. A. Sex Differences in the Relationship between C-Reactive Protein and Body Fat. J Clin Endocrinol Metab 2009, 94 (9), 3251–3258. 10.1210/jc.2008-2406.

(64) Bain, C. R.; Myles, P. S.; Martin, C.; Wallace, S.; Shulman, M. A.; Corcoran, T.; Bellomo, R.; Peyton, P.; Story, D. A.; Leslie, K.; Forbes, A. Postoperative Systemic Inflammation after Major Abdominal Surgery: Patient-centred Outcomes. Anaesthesia 2023, 78 (11), 1365–1375. 10.1111/anae.16104.

(65) Calder, P. C. Fatty Acids and Inflammation: The Cutting Edge between Food and Pharma. European Journal of Pharmacology 2011, 668, S50–S58. 10.1016/j.ejphar.2011.05.085.

(66) Liss, K. H. H.; Lutkewitte, A. J.; Pietka, T.; Finck, B. N.; Franczyk, M.; Yoshino, J.; Klein, S.; Hall, A. M. Metabolic Importance of Adipose Tissue Monoacylglycerol Acyltransferase 1 in Mice and Humans. Journal of Lipid Research 2018, 59 (9), 1630–1639. 10.1194/jlr.M084947.

(67) Gatson, J.; Zang, Q.; Maass, D.; Minei, J.; Idris, A.; Pepe, P.; Wigginton, J. Estrogen Decreases Inflammation and Apoptotic Signaling in the Heart Following Severe Burn Injury. Critical Care 2010, 14 (1), P584. 10.1186/cc8816.

(68) Gatson, J. W.; Maass, D. L.; Simpkins, J. W.; Idris, A. H.; Minei, J. P.; Wigginton, J. G. Estrogen Treatment Following Severe Burn Injury Reduces Brain Inflammation and Apoptotic Signaling. J Neuroinflammation 2009, 6, 30. 10.1186/1742-2094-6-30.

(69) Harding, A. T.; Heaton, N. S. The Impact of Estrogens and Their Receptors on Immunity and Inflammation during Infection. Cancers (Basel) 2022, 14 (4), 909. 10.3390/cancers14040909.

(70) Igarashi, M.; Ayabe, T.; Yamamoto-Hanada, K.; Matsubara, K.; Sasaki, H.; Saito-Abe, M.; Sato, M.; Mise, N.; Ikegami, A.; Shimono, M.; Suga, R.; Ohga, S.; Sanefuji, M.; Oda, M.; Mitsubuchi, H.; Michikawa, T.; Yamazaki, S.; Nakayama, S.; Ohya, Y.; Fukami, M. Female-Dominant Estrogen Production in Healthy Children before Adrenarche. Endocr Connect 2021, 10 (10), 1221–1226. 10.1530/EC-21-0134.

(71) Wickens, N.; van Rensburg, E. J.; de Gouveia Belinelo, P.; Milroy, H.; Martin, L.; Wood, F.; Woolard, A. “It’s a Big Trauma for the Family”: A Qualitative Insight into the Psychological Trauma of Paediatric Burns from the Perspective of Mothers. Burns 2024, 50 (1), 262–274. 10.1016/j.burns.2023.06.014.

(72) Woolard, A.; Hill, N. T. M.; McQueen, M.; Martin, L.; Milroy, H.; Wood, F. M.; Bullman, I.; Lin, A. The Psychological Impact of Paediatric Burn Injuries: A Systematic Review. BMC Public Health 2021, 21 (1), 2281. 10.1186/s12889-021-12296-1.

(73) Allahham, A.; Cooper, M. N.; Fear, M. W.; Martin, L.; Wood, F. M. Quality of Life in Paediatric Burn Patients with Non-Severe Burns. Burns 2023, 49 (1), 220–232. 10.1016/j.burns.2022.03.012.

(74) Gavrilova, Y.; Rooney, E.; Donevant, J.; Ficalora, J.; Sieglein, A.; Kahn, S.; Davidson, T. Sex Differences, Age, and Burn Size Contribute to Risk of PTSD and Depression After Burn Injury. J Burn Care Res 2024, 45 (6), 1444–1453. 10.1093/jbcr/irae092.

(75) Allahham, A.; Cooper, M. N.; Mergelsberg, E.; Fear, M. W.; Martin, L. J.; Wood, F. M. A Comparison of Parent-Reported and Self-Reported Psychosocial Function Scores of the PedsQL for Children with Non-Severe Burn. Burns 2023, 49 (5), 1122–1133. 10.1016/j.burns.2022.09.001.

(76) Miranda, A. M.; Oliveira, T. G. Lipids under Stress – a Lipidomic Approach for the Study of Mood Disorders. BioEssays 2015, 37 (11), 1226–1235. 10.1002/bies.201500070.

(77) Norbury, W. B.; Herndon, D. N.; Branski, L. K.; Chinkes, D. L.; Jeschke, M. G. Urinary Cortisol and Catecholamine Excretion after Burn Injury in Children. J Clin Endocrinol Metab 2008, 93 (4), 1270–1275. 10.1210/jc.2006-2158.

(78) Jeschke, M. G.; Mlcak, R. P.; Finnerty, C. C.; Norbury, W. B.; Przkora, R.; Kulp, G. A.; Gauglitz, G. G.; Zhang, X.-J.; Herndon, D. N. Gender Differences in Pediatric Burn Patients: Does It Make a Difference? Ann Surg 2008, 248 (1), 126–136. 10.1097/SLA.0b013e318176c4b3.

(79) Short, S. J.; Stalder, T.; Marceau, K.; Entringer, S.; Moog, N. K.; Shirtcliff, E. A.; Wadhwa, P. D.; Buss, C. Correspondence between Hair Cortisol Concentrations and 30-Day Integrated Daily Salivary and Weekly Urinary Cortisol Measures. Psychoneuroendocrinology 2016, 71, 12–18. 10.1016/j.psyneuen.2016.05.007.

(80) Prodam, F.; Ricotti, R.; Agarla, V.; Parlamento, S.; Genoni, G.; Balossini, C.; Walker, G. E.; Aimaretti, G.; Bona, G.; Bellone, S. High-End Normal Adrenocorticotropic Hormone and Cortisol Levels Are Associated with Specific Cardiovascular Risk Factors in Pediatric Obesity: A Cross-Sectional Study. BMC Medicine 2013, 11 (1), 44. 10.1186/1741-7015-11-44.

(81) Anni, N. S.; Jung, S. J.; Shim, J.-S.; Jeon, Y. W.; Lee, G. B.; Kim, H. C. Stressful Life Events and Serum Triglyceride Levels: The Cardiovascular and Metabolic Diseases Etiology Research Center Cohort in Korea. Epidemiol Health 2021, 43, e2021042. 10.4178/epih.e2021042.

(82) Egberts, M. R.; Geenen, R.; de Jong, A. E.; Hofland, H. W.; Van Loey, N. E. The Aftermath of Burn Injury from the Child’s Perspective: A Qualitative Study. J Health Psychol 2020, 25 (13–14), 2464–2474. 10.1177/1359105318800826.

(83) Panayi, A. C.; Heyland, D. K.; Stoppe, C.; Jeschke, M. G.; Didzun, O.; Matar, D.; Tapking, C.; Palackic, A.; Bliesener, B.; Harhaus, L.; Knoedler, S.; Haug, V.; Bigdeli, A. K.; Kneser, U.; Orgill, D. P.; Hundeshagen, G. The Long-Term Intercorrelation between Post-Burn Pain, Anxiety, and Depression: A Post Hoc Analysis of the “RE-ENERGIZE” Double-Blind, Randomized, Multicenter Placebo-Controlled Trial. Critical Care 2024, 28 (1), 95. 10.1186/s13054-024-04873-8.

(84) Wasiak, J.; Lee, S. J.; Paul, E.; Shen, A.; Tan, H.; Cleland, H.; Gabbe, B. Female Patients Display Poorer Burn-Specific Quality of Life 12 Months after a Burn Injury. Injury 2017, 48 (1), 87–93. 10.1016/j.injury.2016.07.032.

(85) van Baar, M. E.; Polinder, S.; Essink-Bot, M. L.; van Loey, N. E. E.; Oen, I. M. M. H.; Dokter, J.; Boxma, H.; van Beeck, E. F. Quality of Life after Burns in Childhood (5–15 Years): Children Experience Substantial Problems. Burns 2011, 37 (6), 930–938. 10.1016/j.burns.2011.05.004.

(86) Cuttle, L.; Fear, M.; Wood, F. M.; Kimble, R. M.; Holland, A. J. A. Management of Non-Severe Burn Wounds in Children and Adolescents: Optimising Outcomes through All Stages of the Patient Journey. Lancet Child Adolesc Health 2022, 6 (4), 269–278. 10.1016/S2352-4642(21)00350-3.

(87) Begum, S.; Johnson, B. Z.; Morillon, A.-C.; Yang, R.; Bong, S. H.; Whiley, L.; Gray, N.; Fear, V. S.; Cuttle, L.; Holland, A. J. A.; Nicholson, J. K.; Wood, F. M.; Fear, M. W.; Holmes, E. Systemic Long-Term Metabolic Effects of Acute Non-Severe Paediatric Burn Injury. Sci Rep 2022, 12 (1), 13043. 10.1038/s41598-022-16886-w.

