## Supplementary material for "Sex-Based Differences in Long-Term Lipid Metabolism, Inflammation and Stress Regulation After Non-Severe Paediatric Burns": Figure S1

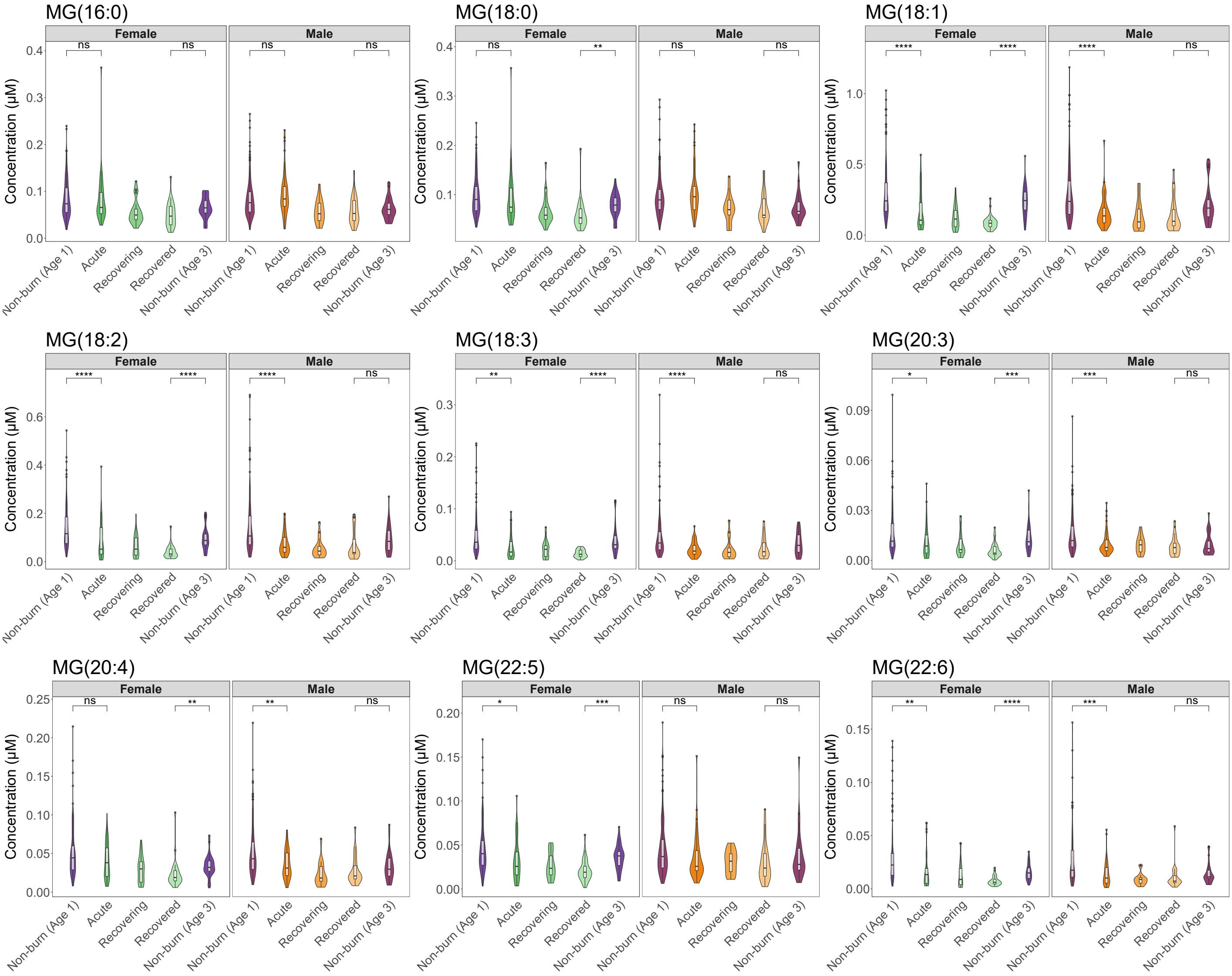


**Figure S1. Monoacylglycerol lipid species detected in the paediatric burn and non-burn cohorts.** Individual lipid species are split by sex (Female – left, Male – right) and concentrations shown with violin plots for non-burn controls at age 1, paediatric burn patients during the 'acute' phase, 'recovering' and 'recovered' stages and non-burn controls at age 3. Significance is denoted by '*' after adjustment with ns = not significant, '*' = *p*-value <0.05, '**' = *p*-value <0.01, '***' = *p*-value <0.001 and '****' = *p*-value <0.0001.
